# Meta-regression of Genome-Wide Association Studies to estimate age-varying genetic effects

**DOI:** 10.1101/2023.01.25.23284845

**Authors:** Panagiota Pagoni, Julian P. T. Higgins, Deborah A. Lawlor, Evie Stergiakouli, Nicole M. Warrington, Tim T. Morris, Kate Tilling

## Abstract

**Background:** Fixed-effect meta-analysis has been used to summarize genetic effects on a phenotype across multiple Genome-Wide Association Studies (GWAS) assuming a common underlying genetic effect. Genetic effects may vary with age, therefore meta-analysing GWAS of age-diverse samples could be misleading. Meta-regression allows adjustment for study specific characteristics and models heterogeneity between studies. The aim of this study was to explore the use of meta-analysis and meta-regression for estimating age-varying genetic effects on phenotypes.

**Methods:** With simulations we compared the performance of meta-regression to fixed-effect and random -effects meta-analyses in estimating (i) main genetic effects and (ii) age-varying genetic effects (SNP by age interactions) from multiple GWAS studies under a range of scenarios. We applied meta-regression on publicly available summary data to estimate the main and age-varying genetic effects of the *FTO* SNP rs9939609 on Body Mass Index (BMI).

**Results:** Fixed-effect and random-effects meta-analyses accurately estimated genetic effects when these did not change with age. Meta-regression accurately estimated both the main genetic effects and the age-varying genetic effects. When the number of studies or the age-diversity between studies was low, meta-regression had limited power. In the applied example, each additional minor allele (A) of rs9939609 was inversely associated with BMI at ages 0 to 3, and positively associated at ages 5.5 to 13. This is similar to the association that has been previously reported by a study that used individual participant data.

**Conclusions:** GWAS using summary statistics from age-diverse samples should consider using meta-regression to explore age-varying genetic effects.

**KEY MESSAGES:**

- Meta-analysis has been used to summarize genetic effects on a phenotype across multiple Genome-Wide Association Studies (GWAS) assuming a common underlying genetic effect for all studies. However, genetic effects may vary with age, therefore meta-analysing GWAS of age-diverse samples could produce misleading results.
- Meta-regression could be used to relate observed between-study heterogeneity to study characteristics such as age. Therefore, meta-regression could be used to combine summary level GWAS data to provide evidence for any age-varying genetic effects.
- This simulation study shows that when genetic effects vary with age, meta-regression provides unbiased estimates of main and age-varying genetic effects. The precision of the estimates depends on the number of studies included, and the diversity in age between them.
- The applied example using publicly available summary data, supported the simulation study.
- By applying meta-regression, we observed a previously reported age-varying association between each additional minor allele (A) of rs9939609 and BMI; an inverse at ages 0 to 3 and a positive association at ages 5.5 to 13.Similar association has been previously reported by a study that used individual participant data.

## INTRODUCTION

Genome–wide association studies (GWAS) test associations of millions of single nucleotide polymorphisms (SNPs) across the genome with a phenotype. As SNP effects are generally small, large sample sizes are required for adequate statistical power. This is commonly achieved through fixed effect meta-analysis of summary genetic effects across several GWAS, which increases sample size and statistical power without sharing individual participant data.

Fixed-effect meta-analysis, which assumes a common true underlying genetic effect for all studies (1), has been favored over random-effects meta-analysis, mostly due to its increased statistical power (2). Fixed-effect meta-analysis ignores heterogeneity of genetic effects between studies, and it has been suggested that this could introduce high rates of false positive and/or false negative findings (2, 3). For example, genetic effects may vary with age (4, 5). Therefore, meta-analysing GWAS studies of age-diverse samples with a fixed-effect model, without considering potential heterogeneity of genetic effects due to age, could fail to identify clinically important changes of genetic risk with age. Moreover, ignoring age-varying genetic effects in GWAS may lead in spurious results in other methods that use GWAS summary data as input to estimate: genetic correlation between traits (LD score regression) (6), genetic predisposition to a trait (Polygenic Risk Scores) (7) and the causal effect of an exposure on an outcome (Two-sample Mendelian randomization) (8).

An approach recommended in meta-analysis of Randomized Controlled Trials (RCTs) to estimate treatment-covariate interactions (e.g., treatment-age interactions) is a two-stage approach, where the interaction is estimated within each study, and these interactions are then meta-analysed (9). This approach would have limited application in GWAS as most studies do not perform or report an interaction analysis (e.g., SNP-age interaction effects).

An alternative method that could be used is meta-regression, which uses summary data and relates observed between-study heterogeneity to study characteristics and investigates the impact of moderator variables on estimated genetic effect sizes (10, 11). Meta-regression has not been widely applied in RCTs due to limited statistical power, related to both the size of the individual studies and the number of studies included (12). A search of the literature and GWAS data bases in July 2022, suggests that meta-regression has been applied in only two GWASs to explore age related differences between included studies. In both cases, genetic variants with age-varying genetic effects were identified (13, 14). We have not identified published research exploring the conditions under which meta-regression outperforms meta-analysis when age-varying genetic effects exist.

The aim of this study was to explore the use of meta-analysis and meta-regression to examine age-varying genetic effects on phenotypes, using summary GWAS data. We compared the performance of meta-regression and fixed-effect and random-effects meta-analysis in estimating (i) main genetic effects (i.e., the effect at age 0) and (ii) age-varying genetic effects (SNP by age interactions) using multiple simulated cross-sectional GWAS studies. We simulated phenotype-genotype associations under a range of data generating processes, varying the number of studies and sample sizes, the overlap in the age range of study participants (i.e., age-diversity), and the sampling variability within and between studies. Subsequently, we applied meta-analysis and meta-regression to estimate the age-varying genetic associations between the rs9939609 SNP at the *FTO* locus and body mass index (BMI) across early life-course, using publicly available summary data, and compare these to estimates from previous individual-participant analyses.

## METHODS

### Data generating mechanisms for simulations

Participant age (*age*_*ij*_ for participant *i* in study *j*), drawn from a uniform distribution, was set between 10 and 59 years. A single SNP with a large effect size, *SNP*_*ij*_, was simulated with a minor allele frequency (MAF) of 0.2 and the number of risk alleles (0,1,2) was drawn from a binomial distribution. We generated the outcome phenotype (*Y*_*ij*_) to be dependent on: *Scenario 1*. age and genotype; *Scenario 2*. age and genotype, with an interaction between age and genotype (linear interaction term); *Scenario 3*. age, genotype and a quadratic term of age; *Scenario 4*. genotype, age and a quadratic term of age, with an interaction between age and genotype; *Scenario 5*. genotype, age and a quadratic term of age, where genotype interacts with age and quadratic age (non-linear interaction term). Equations for the phenotype generating scenarios and the parameter values are presented in Table 1. We assumed that the effect of age on phenotype (β_*age*_) was identical for each study but that the effect of genotype varied randomly across studies (*β*_*SNP*_ µ µ_*j*_), corresponding to a random-effects meta-analysis model for the genotype-phenotype association. As a “base case” scenario, we used 1SD within and between study variability (ε_*ij*_~*N*(0,1) and µ_*j*_~*N*(0,1)) in the data generating mechanisms, with 40 cross-sectional studies each with sample size *N*_*j*_ = 1,000.

**Table 1.** Equations underlying the phenotype generating mechanism for each simulated scenario and parameter values for ‘Base case scenario’.

| Simulated scenarios |  |  |
| --- | --- | --- |
| Scenario 1 | $Y_{ij} = \beta_0 + \beta_{age} \times age_{ij} + (\beta_{SNP} + u_j) \times SNP_{ij} + \varepsilon_{ij}$ | |
| Scenario 2 | $Y_{ij} = \beta_0 + \beta_{age} \times age_{ij} + (\beta_{SNP} + u_j) \times SNP_{ij} + \beta_{SNP \times age} \times age_{ij} \times SNP_{ij} + \varepsilon_{ij}$ | |
| Scenario 3 | $Y_{ij} = \beta_0 + \beta_{age} \times age_{ij} + \beta_{age^2} \times age_{ij}^2 + (\beta_{SNP} + u_j) \times SNP_{ij} + \varepsilon_{ij}$ | |
| Scenario 4 | $Y_{ij} = \beta_0 + \beta_{age} \times age_{ij} + \beta_{age^2} \times age_{ij}^2 + (\beta_{SNP} + u_j) \times SNP_{ij} + \beta_{SNP \times age} \times age_{ij} \times SNP_{ij} + \varepsilon_{ij}$ | |
| Scenario 5 | $Y_{ij} = \beta_0 + \beta_{age} \times age_{ij} + \beta_{age^2} \times age_{ij}^2 + (\beta_{SNP} + u_j) \times SNP_{ij} + \beta_{SNP \times age} \times age_{ij} \times SNP_{ij} + \beta_{SNP \times age^2} \times age_{ij}^2 \times SNP_{ij} + \varepsilon_{ij}$ | |
| Parameter | Value | Interpretation |
| $\beta_0$ | 25 | Baseline mean value of phenotype when $age_{ij} = 0$ and $SNP_{ij} = 0$ |
| $\beta_{age}$ | 0.010 | Effect of age on phenotype |
| $age_{ij}$ | $\sim U(\min age, \max age)$ | Age of participant $i$ in study $j$ |
| $\beta_{SNP}$ | 1.5 | Effect of genetic variant on phenotype (main genetic effect) |
| $SNP_{ij}$ | 0,1,2 | Number of alleles of a genetic variant for participant $i$ in study $j$ |
| $\beta_{SNP \times age}$ | 0.020 | Age varying-genetic effect on phenotype (linear interaction term) |
| $\beta_{SNP \times age^2}$ | 0.001 | Non-linear age-varying genetic effect on phenotype (non-linear interaction term) |
| $\beta_{age^2}$ | 0.001 | Non-linear effect of age on phenotype |
| $\varepsilon_{ij}$ | $\sim N(0,1)$ | Within study sampling error |
| $u_j$ | $\sim N(0,1)$ | Between study sampling error |
| *j*th study $j = (1, 2, \dots, 40)$, *i*th participant $i = (1, 2, \dots, 1000)$ | | |

### Estimating study-specific genotype-phenotype associations

Within each cross-sectional study, we used linear regression to estimate the genotype-phenotype association. As is usual in GWAS studies, models were adjusted only for age, and no further adjustments were made to account for non-linearity and SNP-age interactions. Equation *(1)* describes the regression models:

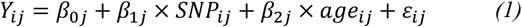

We collected the estimated genotype-phenotype effect estimate 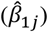 and its standard error 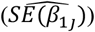 from each study, in addition to the mean age 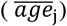 of participants in each study.

### Description of compared methods

#### Meta-analysis

Fixed-effect meta-analysis assumes that all studies draw a (random) sample from the same underlying population and hence share a common true effect size for each SNP. The pooled meta-analysis estimates the population average effect (15). The estimated effect for a given SNP in each study is:

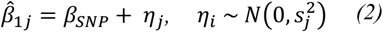

where, 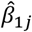 the genotype-phenotype effect in the jth study, *β*_*SNP*_ is the common genetic effect, and η_*j*_ is random error describing the sampling variability within each study (with variance 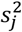in study *j*, i.e., the variance of 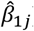).

Random-effects meta-analysis allows the true genetic effect size to differ across studies. Here,

*β*_*SNP*_ reflects an estimate of the average effect across study populations. The estimated effect for a given SNP in each study is:

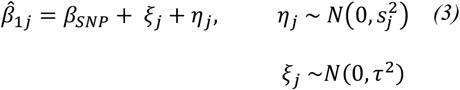

where, *β*_*SNP*_ is the mean genetic effect, ξ_*j*_ represents heterogeneity, i.e. the study-specific deviation from the mean genetic effect (with variance τ^2^ across studies, i.e. the between study variability), and η_*j*_ is random error describing the sampling variability within each study (with variance 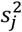in study *j*,i.e. the variance of 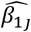). Further information about the estimation of combined genetic effects in fixed-effect and random-effects meta-analysis can be found in Supplementary Note S1. To estimate the between-study variance τ^2^, we used restricted maximum likelihood (REML) method (16).

#### Meta-regression

Random-effects meta-regression extends the random-effects meta-analysis model as follows:

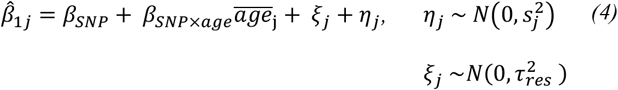

and could also be further extended to include non-linear terms such as:

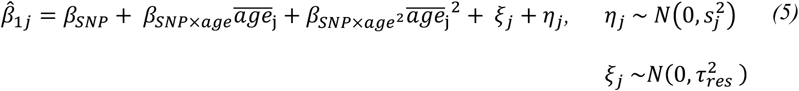

where, β_*SNP*×*age*_ is the difference in the mean effect of a given SNP for each one year increase in age, β_*SNP*×*age*_^2^ is the difference in the mean effect of a given SNP for each one year difference in the square of age and 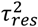 is the residual heterogeneity after accounting for the age effect(s). Meta-regression estimates these two parameters 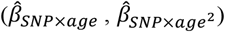 and an intercept term 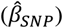 representing the effect of genotype on phenotype for age = 0 (referred to as the main genetic effect). To estimate the between-study variance 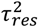we used REML (16).

#### Implementation

For each scenario, we ran 1,000 iterations. We varied i) study sample sizes from 1,000 to 10,000, ii) the number of studies from 10 to 80 and iii) the overlap of age distributions across studies (i.e., age-diversity) from no overlap (0%) to complete overlap (100%) in 25% increments. Figure 1 depicts the overlap of age distributions between studies and further information about the age ranges of each study can be found in the Table S1. Lastly, iv) we varied the within and between study variability from 1 (ε_*ij*_~*N*(0,1) and µ_*j*_~*N*(0,1)) to 3 (ε_*ij*_~*N*(0,3^2^) and µ_*j*_~*N*(0,3^2^)).

**Figure 1.**
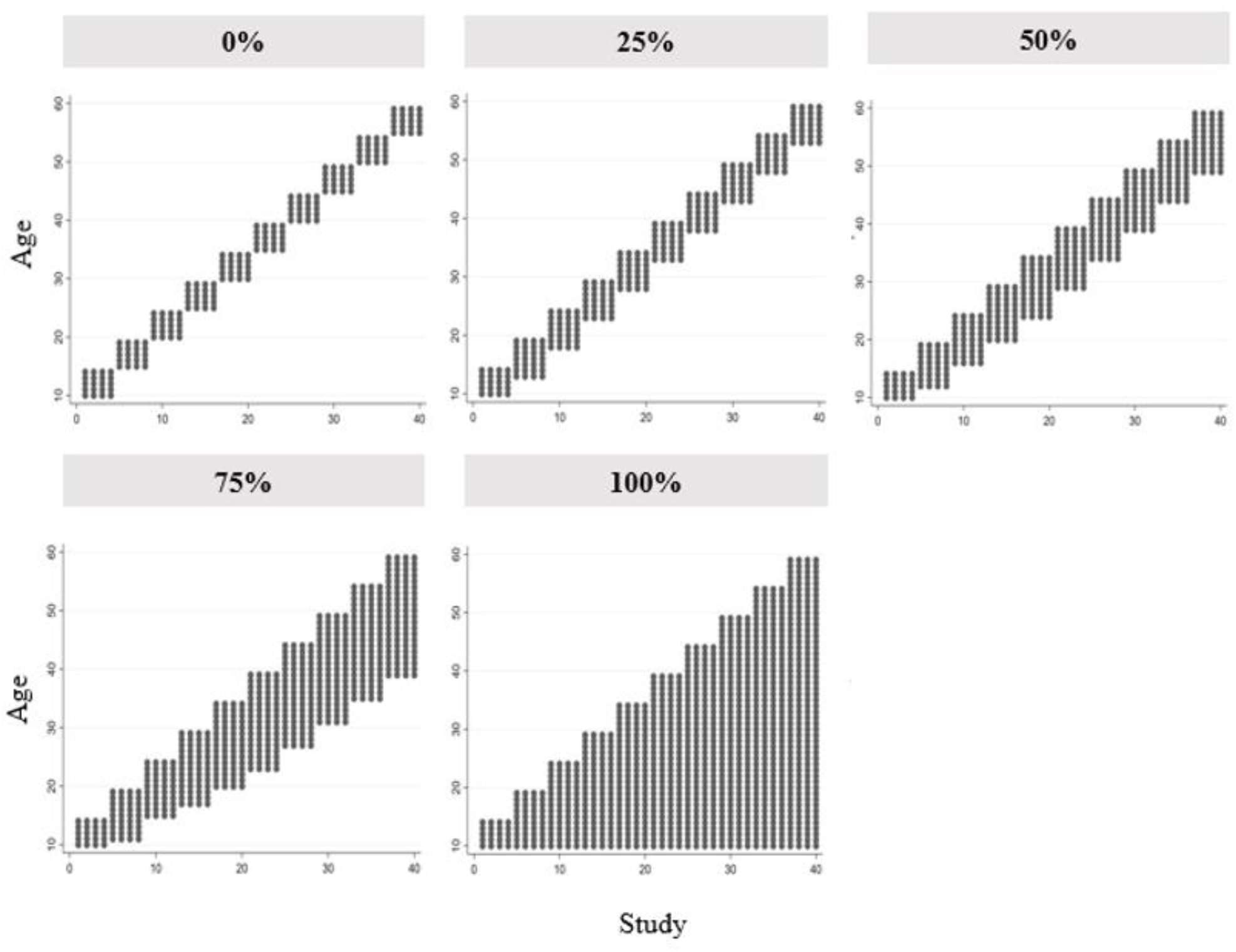
Scatter plot of age of each participant within each study to show age overlap.

#### Estimands and performance measures

The estimands of interest were the main genetic effect (β_SNP_) (the effect if the population had mean age=0), the linear age-varying genetic effect (β_SNP×Age_), the non-linear age-varying effect (β_SNP×*Age*_^2^), and the standard errors (SE) of these parameters across simulations.

We present five performance measures: the mean estimate, the bias (the deviation of the estimated parameter from the simulated value), the coverage of the 95% confidence interval (CI) (the proportion of simulated datasets for which the 95% confidence interval included the simulated value), the empirical standard error (Emp SE), and the mean standard error (Mean SE).

#### Estimating the age-varying genetic association between the rs9939609 SNP at the *FTO* locus and body mass index (BMI)

We used a real data example to illustrate the application of meta-analysis and meta-regression in estimating age-varying genetic effects using summary level association statistics. An age-varying association between the rs9939609 SNP at the *FTO* locus and body mass index (BMI) has been previously demonstrated (4, 5, 17, 18). We extracted summary level data for the association between rs9939609 and BMI from a study investigating the effect of this genetic variant on BMI from infancy to late childhood (5). The effect of rs9939609 on BMI was estimated in 8 cohorts (N=569 to 7,482) at up to 10 ages within each cohort from 0 to 13 years. Detailed information about effect sizes within each cohort at each time point can be found in Supplementary Table S2.

We estimated the association between rs9939609 SNP and BMI using fixed-effect meta-analysis and meta-regression adjusting for a cubic term of age, which can be written as follows:

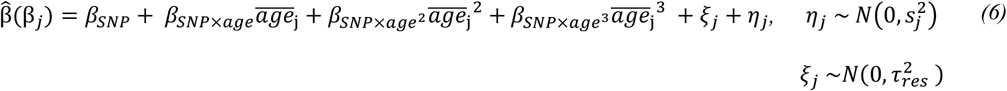

The choice to adjust for a cubic term of age was made based on evidence suggesting that each additional minor allele (A) of this variant is inversely associated with BMI from ages 0 to 3 and positively associated from ages 5.5 to 13 (5). Effect sizes were estimated at multiple time points within the same cohorts, so we used generalized weights to adjust standard errors for the sample overlap (19).

## RESULTS

### Scenarios 1 & 3: Data generated with no age-varying genetic effect

#### i) Estimation of main genetic effect (β_SNP_) (i.e., the effect in a population with mean age=0)

As expected, as there was no age-varying genetic effect, both the fixed-effect and the random effects meta-analyses yielded unbiased estimates of the main genetic effect (β_SNP_) across all proportions of overlapping ages between simulated studies (Figure 2 A & B), although CI coverage was below the nominal 95% level for the fixed-effect meta-analysis. Similarly, meta-regression models including either a linear or a quadratic term of age demonstrated negligible bias for the main genetic effect (Figure 2 C & D), but CI coverage was slightly below the nominal level for models including a quadratic term of age (Table S3).

**Figure 2.**
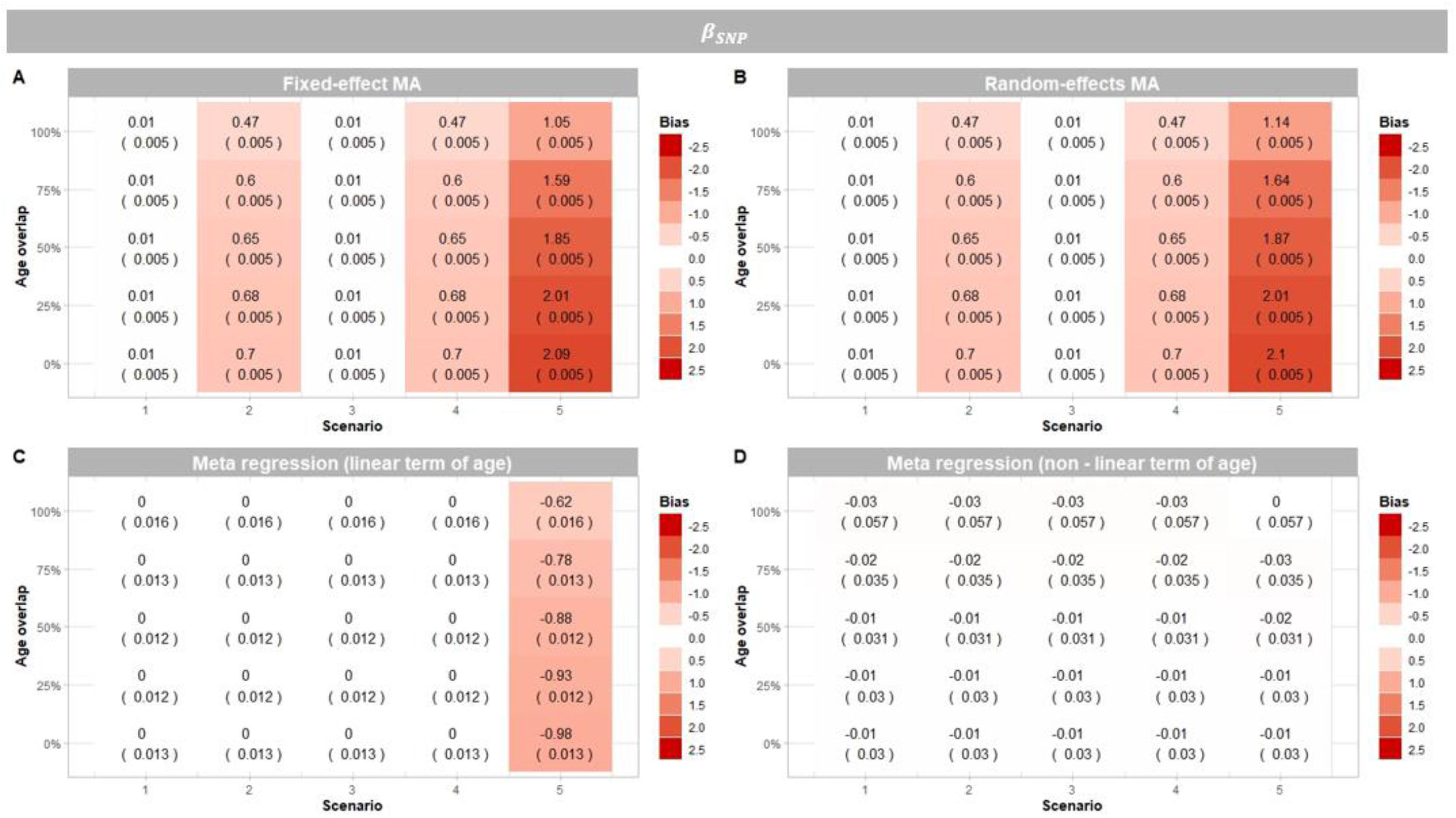
Heat maps displaying the absolute bias (Monte Carlo standard error) for each method, scenario and age overlap for the main genetic effect (i.e., the genetic effect in a population of mean age=0) (*β*_*SNP*_) (N =1,000). MA: Meta-analysis

#### ii) Estimation of age-varying genetic effects (β_SNP×Age_, β_SNP×Age_^²^)

Both meta-regression models (including a linear or quadratic term of age) yielded unbiased (i.e., mean of zero) estimates of the linear age-varying genetic effect (β_SNP×Age_) (Figure 3A & B) and the non-linear age-varying genetic effect (β_SNP×Age_^2^) (Figure 3C). However, for both estimands (β_SNP×Age_, β_SNP×Age_^2^), the values estimated by the meta-regression models (erroneously including a linear or a quadratic term of age) were highly variable as seen by the large Monte Carlo SEs of bias in (Figure 3A-C). As the proportion of overlapping ages between simulated studies was increased, the variability of estimated values increased.

**Figure 3.**
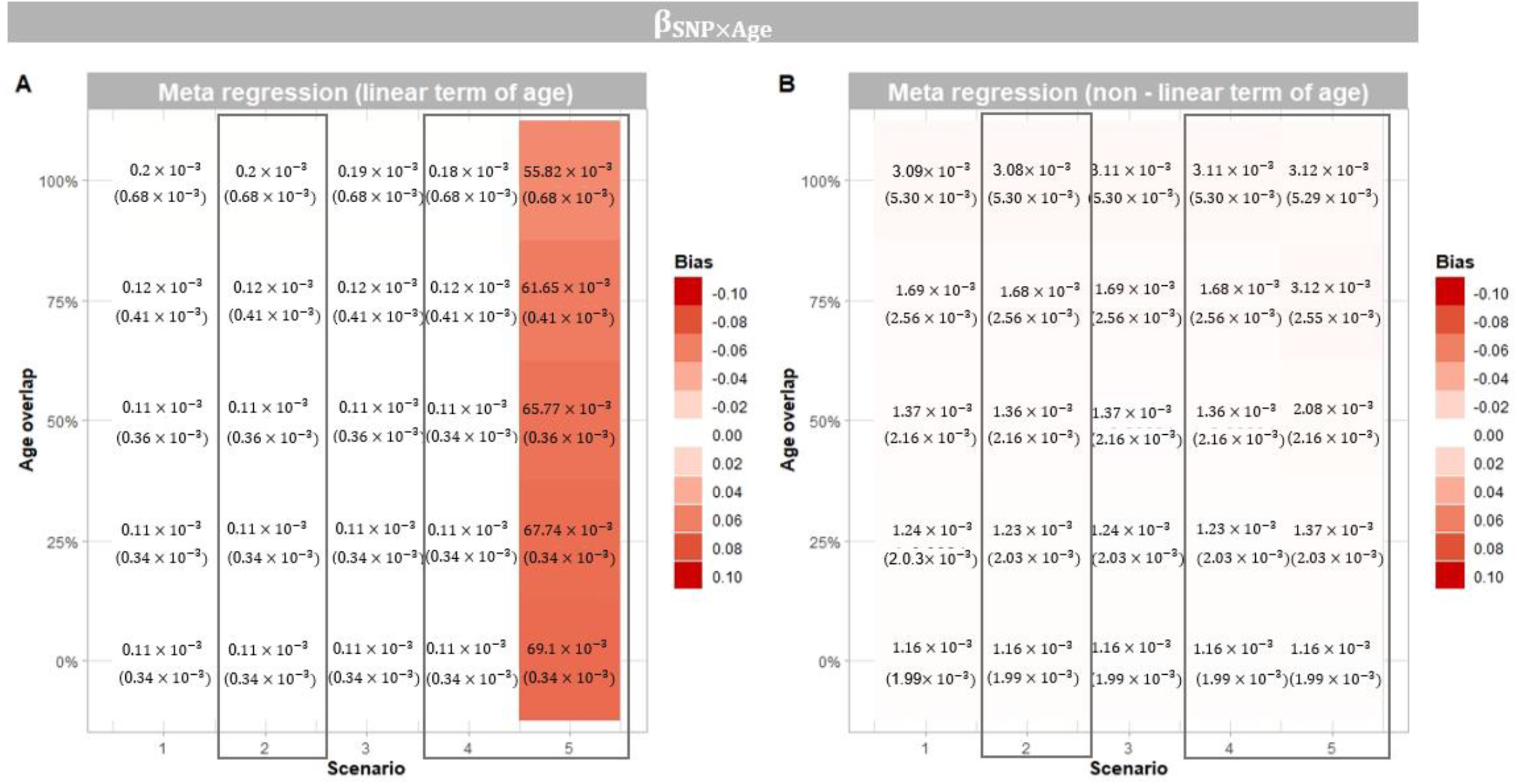
**A-B**. Heat maps displaying the absolute bias (Monte Carlo standard error) for each method, scenario and age overlap for the linear and non-linear age-varying genetic effect (β_SNP×Age_, β_SNP×*Age*_^2^) (N =1,000). Squares represent scenarios where β_SNP×Age_ = 0.02 and β_SNP×*Age*_^2^ = 0.001. In other scenarios β_SNP×Age_ = 0 and β_SNP×*Age*_^2^ = 0.

**Figure 3C.**
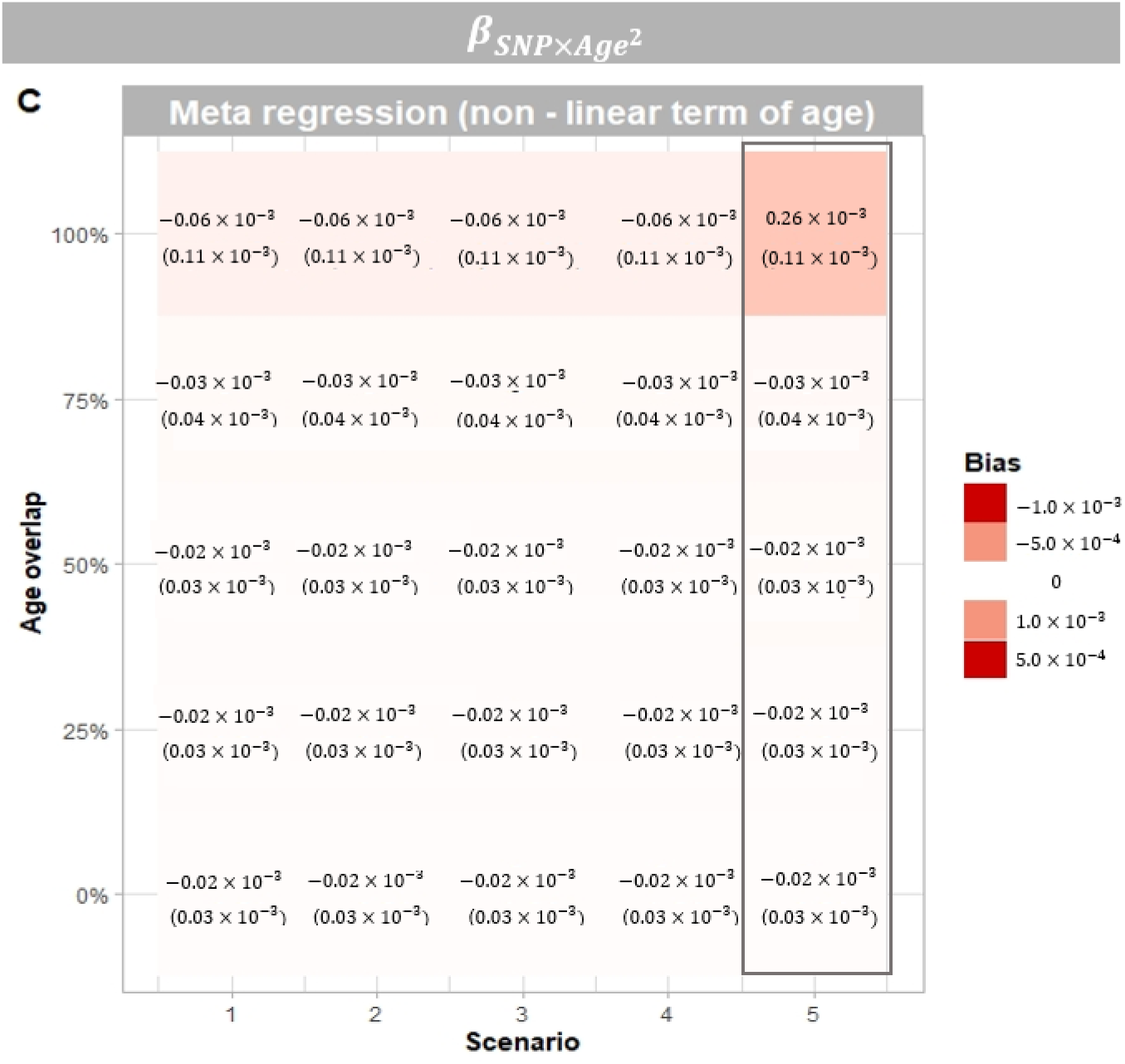
Heat maps displaying the absolute bias (Monte Carlo standard error) for each method, scenario and age overlap for the linear and non-linear age-varying genetic effect (β_SNP×Age_, β_SNP×*Age*_^2^) (N =1,000). Squares represent scenarios where β_SNP×Age_ = 0.02 and β_SNP×*Age*_^2^ = 0.001. In other scenarios β_SNP×Age_ = 0 and β_SNP×*Age*_^2^ = 0.

CI coverage for the linear age-varying genetic effect (β_SNP×Age_ = 0) was consistent with the nominal 95% level for meta-regression models including a linear term of age, CI coverage was below the nominal 95% level for the linear and non-linear age-varying genetic effect (β_SNP×Age_, β_SNP×Age_^2^) in meta-regression models including a quadratic term of age (Table S4 & S5).

### Scenarios 2 & 4: Data generated with a linear age-varying genetic effect

#### i) Estimation of main genetic effect (β_SNP_) (i.e., the effect in a population with mean age=0)

When there were linear age-varying genetic effects, both fixed and random-effects meta-analyses gave biased estimates of the main genetic effect, across all proportions of overlapping ages between simulated studies (Figure 2 A & B). Meta-regression models including a linear or quadratic term of age produced unbiased estimates of the main genetic effect (Figure 2 C & D). As the age overlap between studies increased, the meta-regression estimates were more variable, as seen by the large Monte Carlo SEs.

In both scenarios, CI coverage for the main genetic effect (i.e., the effect if the population had a mean age of 0) was consistently below the nominal 95% level for fixed-effect and random-effects meta-analyses, across all proportions of overlapping ages between simulated studies. Meta-regression including a linear term of age yielded coverage of CIs consistent with the nominal 95% level. When the meta-regression included a quadratic term of age, coverage of CIs was consistently below the nominal 95% level (Table S3).

#### ii) Estimation of age-varying genetic effect (β_SNP×Age_, β_SNP×Age_^²^)

Meta-regression estimates of the linear age-varying genetic effect (β_SNP×Age_) were unbiased in both meta-regression models (Figure 3A & B). Similarly, estimates of the non-linear age-varying genetic effect (β_SNP×Age_^2^ = 0) was unbiased in the meta-regression model including a quadratic term of age (Figure 3C). The variance of the estimated values increased as the proportion of overlapping ages between simulated studies increased.

In both scenarios, CI coverage for the linear and non-linear age-varying genetic effects (β_SNP×Age_, β_SNP×Age_^2^) were consistent with the nominal 95% level for meta-regression models including a linear term of age but not for meta-regression including a quadratic term of age (Table S4 & S5).

### Scenario 5: Data generated with a quadratic age-varying genetic effect

#### i) Estimation of main genetic effect (β_SNP_) (i.e., the effect in a population with mean age=0)

Both fixed and random meta-analyses gave biased estimates of the main genetic (Figure 2 A & B). Meta-regression with only a linear age term also gave biased estimates of the main genetic effect (Figure 2C). Meta-regression including a quadratic term of age yielded unbiased estimates, but variability of bias increased as age overlaps between studies increased (Figure 2D).

CI coverage for the main genetic effect (i.e., the effect in a population of mean age=0) was consistently below the nominal 95% level for all compared methods, across all proportions of overlapping ages between simulated studies. Meta-regression models including a quadratic term of age demonstrated the highest CI coverage (Table S3).

#### ii) Estimation of age-varying genetic effect (β_SNP×Age_, β_SNP×Age_^²^)

Meta-regression models including only a linear term of age gave biased estimates of the linear age-varying genetic effect (β_SNP×Age_), across all proportions of overlapping ages between simulated studies. In contrast, the meta-regression model also including a quadratic term of age yielded unbiased estimates of both the linear and non-linear age-varying genetic effects (β_*SNP*×*Age*_), (β_SNP×Age_^2^), but variance of estimates increased as proportions of overlapping ages between studies increased (Figure 3A-C).

CI coverage for the linear and non-linear age-varying genetic effect (β_SNP×Age_, β_SNP×Age_^2^) was consistently slightly below the nominal 95% level in both meta-regression models (Table S4 & S5).

#### Comparison of Empirical SE and Mean SE

Across all scenarios and proportions of overlapping ages between studies and for all estimands of interest, the random-effects meta-analysis and meta-regression including both a linear term and quadratic term of age yielded comparable empirical and mean SEs (Table S3-S5). In contrast, the fixed-effect meta-analysis produced mean SEs that were smaller than the empirical SEs, highlighting the incompatibility of fixed-effect meta-analysis to our data-generating mechanisms.

The random-effects meta-analysis and meta-regression (both including a linear and quadratic term of age) produced large mean SEs of the main genetic effect (*β*_*SNP*_). Additionally, including high age-diverse (no (0%) age overlaps in age ranges) studies in the meta-regression models produced more precise estimates of the main genetic effect compared to inclusion of low age-diverse (100% overlap in age ranges) studies (Figure 4).

**Figure 4.**
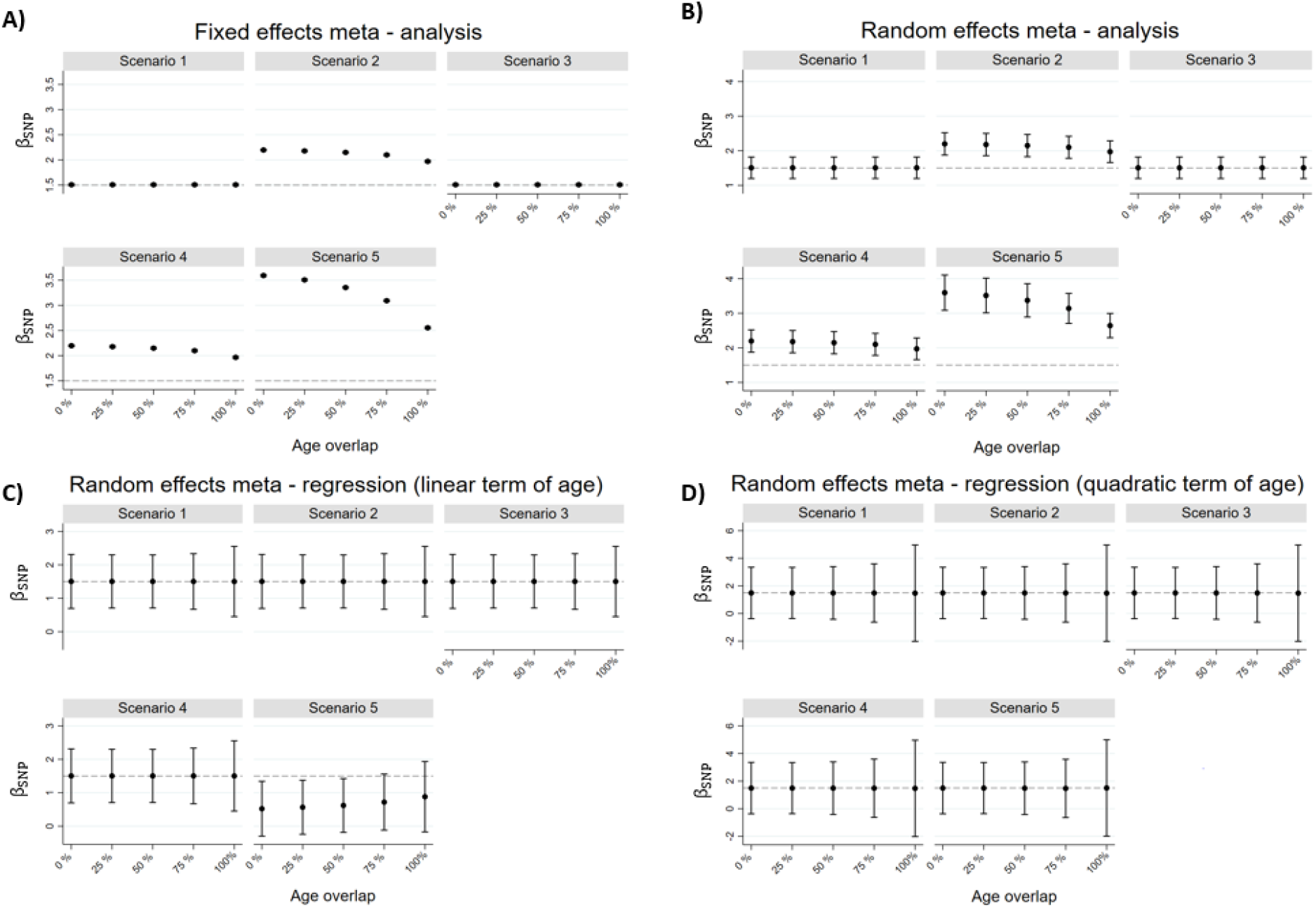
Estimated main genetic effect (i.e., the genetic effect in a population of mean age=0) (*β*_*SNP*_) and 95% confidence intervals. (A) Fixed-effect meta-analysis (B) random-effects meta-analysis, (C) Random-effects meta-regression including a linear term of age (C) Random-effects meta-regression including a quadratic term of age.

The meta-regression models including a linear term of age produced smaller mean SEs for both the linear and non-linear age-varying effects (β_SNP×Age_, β_SNP×Age_^2^) compared with meta-regression models including a quadratic term of age, as expected due to having fewer number of parameters estimated (Figure 5). Moreover, including high age-diverse (no (0%) age overlaps in age ranges) studies in the meta-regression models produced more precise estimates of the linear and non-linear age-varying effects compared to inclusion of low age-diverse (100% overlap in age ranges) studies.

**Figure 5.**
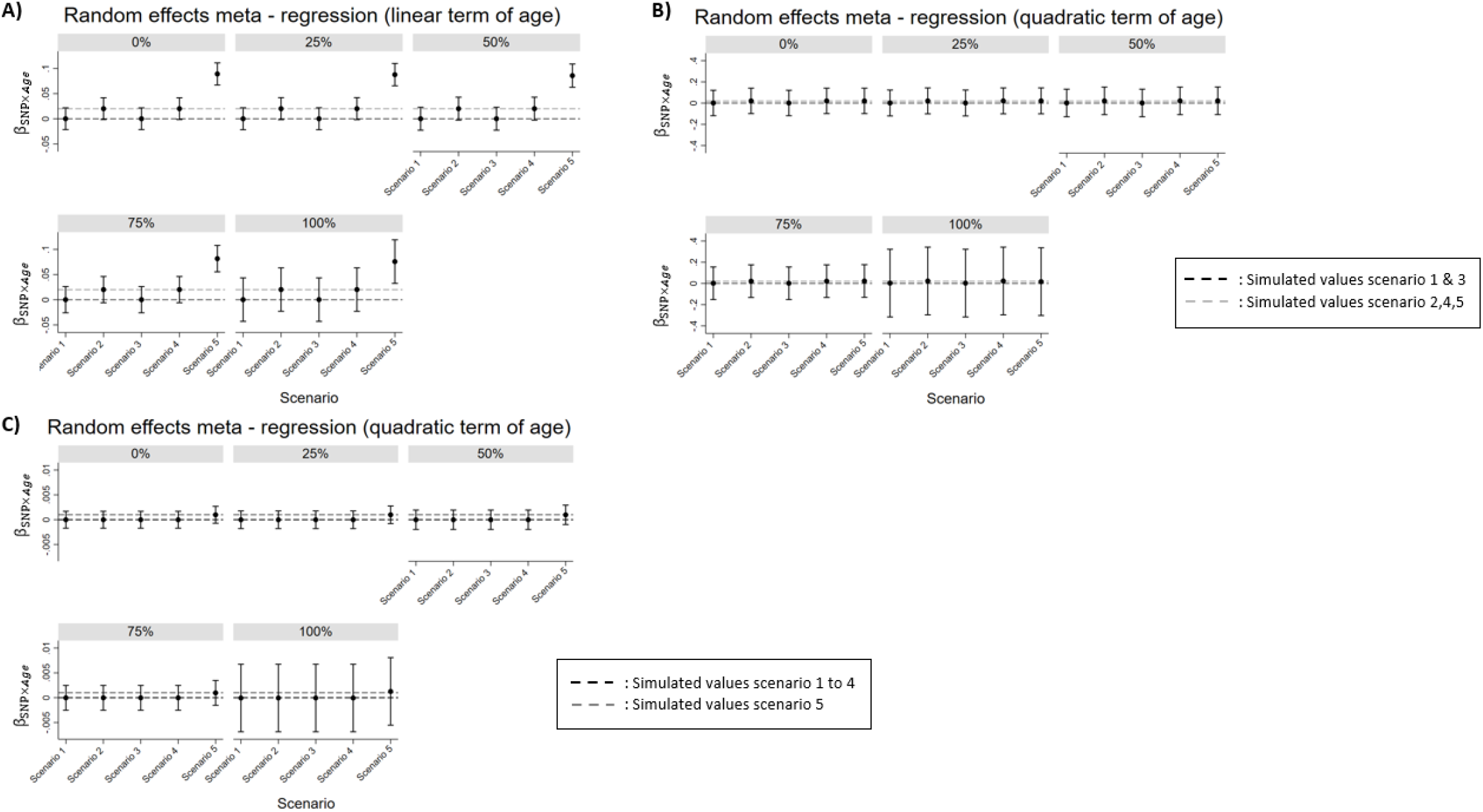
(A) and (B) show the estimated linear age-varying genetic effects (β_SNP×Age_) and their 95% confidence intervals using meta-regression including a linear and a quadratic term by age overlap between studies, respectively. (C) shows the estimated non-linear age-varying genetic effects (β_SNP×*Age*_^2^) and their 95% confidence intervals using meta-regression including a quadratic term by age overlap between studies.

#### Influence of study characteristics

The Supplementary Material (Table S6 – S26) shows results from simulations varying the number of studies included in the analysis (from 10 to 80), sample sizes of each cohort (from 1,000 to 10,000), study level error (µ_*j*_) and individual level error (ε_*ij*_). A small number of studies included in the meta-regression models (including a linear or quadratic term of age) resulted in CI coverage below the nominal 95% level for all estimands of interest (*β*_*SNP*_,β_SNP×Age_, β_SNP×Age_^2^), even when the meta-regression models correctly reflected the data generating mechanisms. Increasing the number of participants within each cohort resulted in decreased mean SEs in fixed effect meta-analysis, while the results remained similar in random-effects methods. As expected, increasing study-level variability resulted in increased mean SEs in all random-random effects methods, while mean SEs in fixed-effect meta-analysis remained unchanged. Conversely, increasing individual-level variability resulted in increased mean SEs in fixed effect meta-analysis and SEs remained unaffected in random-effects methods.

#### Estimating the age-varying genetic association between the rs9939609 SNP at the *FTO* locus and body mass index (BMI)

Detailed information about the effect sizes used in the meta-analysis and meta-regression can be found in Supplementary Table S2. When we applied fixed-effect meta-analysis, a constant negative association (β= -0.05, 95%CI: -0.06 to -0.03) between each additional minor allele (A) of rs9939609 and BMI was estimated. As fixed-effect meta-analysis is a weighted average of all studies, the estimated genetic effect is highly influenced by the fact that most of the largest studies are in early ages, therefore if a different selection of studies was used a different effect may have been estimated. In contrast, when we applied meta-regression, we observed an age-varying association: each additional minor allele (A) of rs9939609 was inversely associated with BMI at ages 0 to 3, and positively associated with BMI at ages 5.5 to 13 (Figure 6). The percentage of variation between studies that can be attributed to heterogeneity rather than chance in fixed-effect meta-analysis was substantial (I^2^ = 76.9%), while adjusting for cubic age using meta-regression reduced the between study heterogeneity (I^2^ = 28.3%) (Table S27). Lastly, the association estimated using meta-regression was similar to the association described in the study we extracted summary data from (5). In that study, individual participant data were utilised to model the median BMI curves of each genotype using the LMS method, and it was observed that carriers of minor alleles (A) showed lower BMI in infancy and higher in childhood.

**Figure 6.**
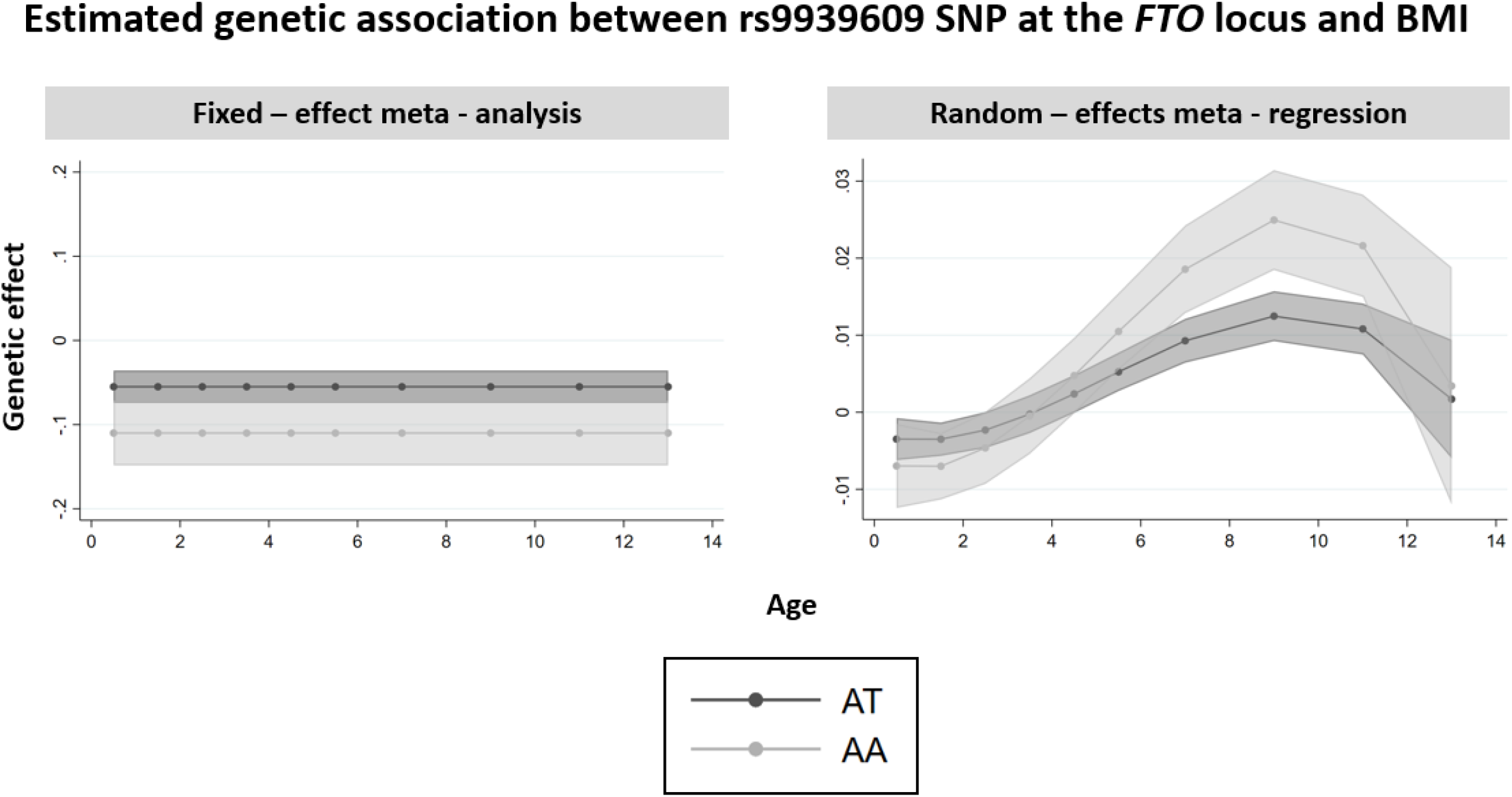
Estimated genetic association between rs9939609 SNP at the *FTO* locus and BMI, as estimated using fixed-effect meta-analysis and meta-regression adjusting for cubic term of age. Number of studies=8, N=19,725

## DISCUSSION

In this study, we compared the performance of meta-regression and meta-analysis in accurately estimating main and age-varying genetic effects (i.e., SNP-age interactions) from simulated and real cross-sectional GWAS studies. Our results demonstrated that fixed-effect and random-effects meta-analyses accurately estimate genetic effects when these are not moderated by age but not when age-varying genetic effects exist. This is because when there is age-moderation of genetic effects, the fixed or random-effects meta-analyses estimate the average effect across the (weighted) age distribution of the studies included, and these estimates are heavily influenced by the amount of data included at each age. In contrast, meta-regression produces unbiased estimates of both the main genetic effects and the age-varying genetic effects, regardless of whether age is a moderator or not. For example, in our real data analysis meta-analysis suggested an inverse association in children aged 0 to 13 years, whereas meta-regression correctly revealed an inverse association in early childhood (0 to 3 years),with this changing to a positive association between age 5.5 and 13 years. However, applying meta-regression when there are no age-varying genetic effects will produce less precise estimates, as more parameters will be estimated.

Exploring age-varying genetic effects in GWAS is important for various reasons. Firstly, it could help to better characterise the extent to which genetic variants which have already been associated with traits in a cross-sectional GWAS also influence change in that phenotype over time. For example, the FTO gene has been consistently reported to be associated with BMI and adiposity related traits, and there is evidence to suggest that this association may be time dependent (17, 18). More specifically, a longitudinal cohort study reports association of the FTO gene with BMI during childhood and up to 20 years of age, when this association starts to get weaker with increasing age (4). Secondly, it could contribute **t**o identifying novel genetic variants, which may be associated with traits only in specific time periods during the life course. For instance, the LEPR locus has been associated with BMI in infancy and it is not linked with adult BMI, suggesting that its effect is no longer present in adulthood (20, 21). Therefore, exploring age-varying genetic effects could contribute to identifying novel genetic variants associated with age of onset, development of traits over time, and disease progression. Thirdly, the increased number of GWAS and the public availability of their results, has increased the popularity of two-sample MR studies, where the effect estimates of the genetic variants of exposure and outcome are extracted from different GWAS (8), allowing estimation of causal effects without requiring the exposure and outcome to be measured in the same participants. The causal effects estimated by MR are often interpreted as “lifetime causal effects of exposures on outcomes”. This interpretation has recently been challenged. More specifically, when the association between genetic variants and exposure is time-varying, then the estimated causal effect might not reflect the lifetime causal effect (22). Therefore, exploring and accurately estimating age-varying genetic effects could help in better characterising causal relationships when the exposure of interest is time-varying.

Even though GWAS commonly include age-diverse samples, meta-regression is rarely used to explore the differences in SNP-phenotype associations due to age. We have identified only two studies that applied meta-regression to account for heterogeneity introduced due to age. A GWAS of bone mineral density (N=30 studies with a total 66,628 participants) applied meta-regression by stratifying the participants in each study into subgroups based on age and adjusting for the median age of each subgroup (13). Two loci (in *ESR1* and *RANKL*) demonstrated age-varying genetic effects, with stronger associations in older age groups. A GWAS of blood pressure traits (N=9 studies and 55,796 participants) applied meta-regression and adjusted for median age of each contributing study; it identified 9 genetic variants with age-varying effects. SNPs located in *CASZ1, EHBP1L1*, and *GOSR2*, demonstrated the largest age-dependent effects, with the effect alleles increasing blood pressure traits in the younger ages and decreasing them in the older (14).

Meta-regression offers a feasible analytic tool to estimate age-varying genetic effects in the framework of GWAS. Similar to many statistical methods, clear research question and justification for applying meta-regression is necessary a priori and careful consideration must be given regarding the data needed. Meta-regression requires only summary level data for the effect of each SNP on the outcome within each study and the median/mean age of participants in each study. As many GWAS on various traits and diseases have already been published and their summary level data are often publicly available, meta-regression maximizes the value of already existing studies to explore age-varying genetic effects. However, it is very often the case that most GWAS consortia provide summary level data of each SNP across studies, but often not by study. For example, in our applied example we originally planned to use publicly available summary data from GWAS consortia but were unable to find any that provided summary data by study. Future GWAS should therefore aim to publish study specific summary results and information about the median/mean age as well as the age range of participants to enable meta-regression. Moreover, our simulation study suggests that consideration should be given to the number of studies and the age-diversity between the studies included in the meta-regression. In Figure 7, we provide guidance regarding these two parameters. When the number of studies included in the meta-regression is low and the age-diversity between samples low, meta-regression has limited power to estimate age-varying genetic effects. Therefore, researchers will need to either include more studies of age-diverse samples or estimate age-varying genetic effects using longitudinal studies. However, when the number of studies included in the meta-regression is high and the age-diversity moderate to high, then meta-regression should be considered as the main analytical approach in GWAS. Lastly, it is important to carefully select whether the application of a linear or non-linear meta-regression is appropriate, as over-misspecification of the model could lead to below nominal CI coverage and under-specification could lead to biased estimates. Information about the effect of a SNP on a phenotype, in relation to age, can be obtained by smaller longitudinal studies, where this relationship can be investigated.

**Figure 7.**
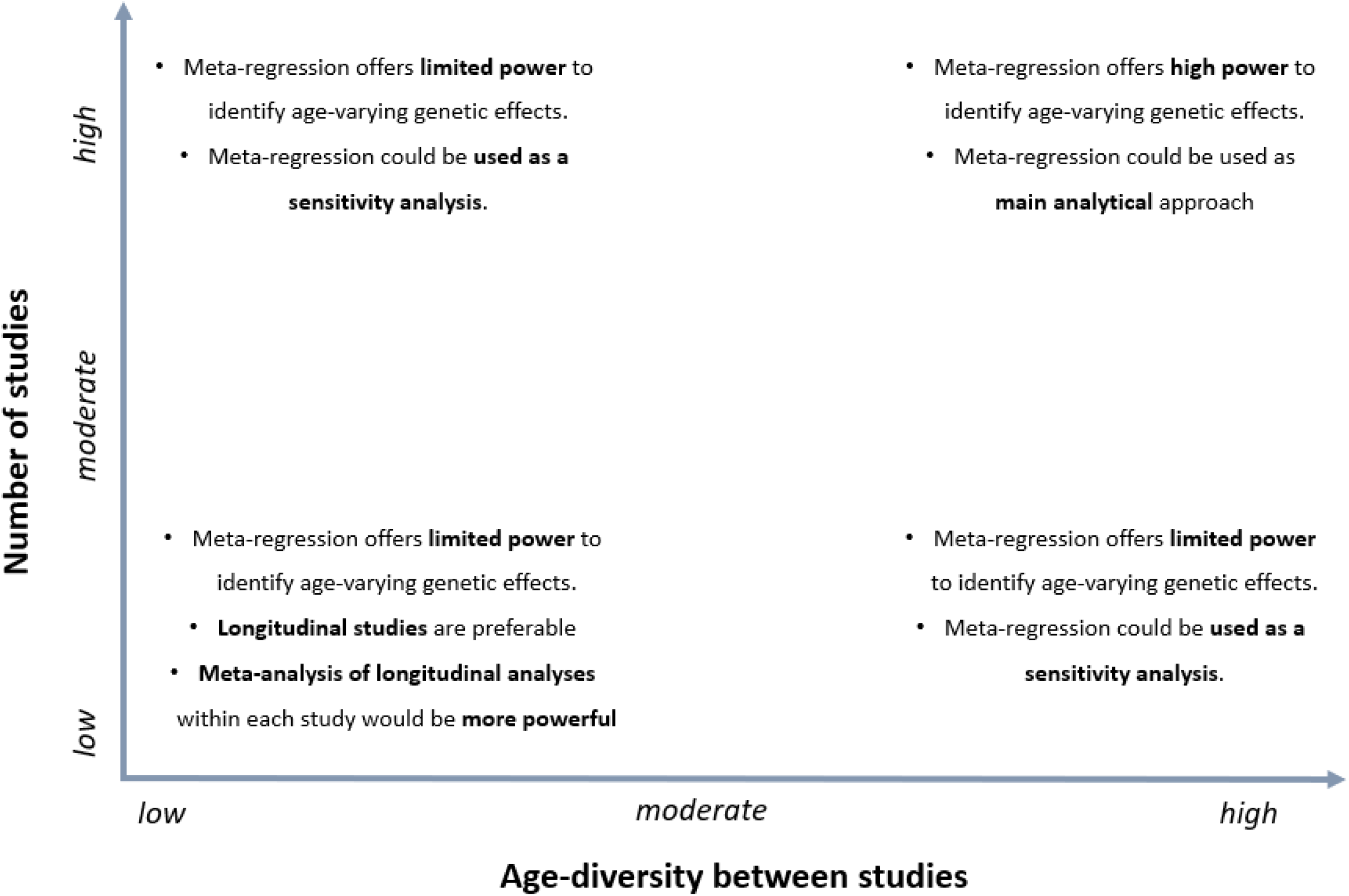
Recommendations for the application of meta-regression in estimating age-varying genetic effects in GWAS, based on the number of studies included and the age-diversity between studies.

Our study has limitations that should be considered. Even though we explored a wide range of plausible scenarios in our simulations, we have inevitably not explored all possible real-world scenarios. For example, further work would be needed to investigate the applicability of our results in cases where the trait of interest is binary/categorical or in cases where the sample size of studies differs and this differentiation is age related (e.g., smaller sample sizes in studies with older participants compared to studies with younger participants). Additionally, we have only investigated the applicability of meta-regression in estimating the association between the genetic effects and quadratic function of age (non-linear age-varying genetic effects). Meta-regression could be easily extended to accommodate higher degree polynomials and splines.

## CONCLUSIONS

Fixed-effect and random-effects meta-analysis that are typically used to synthesize genetic effects from multiple GWAS produce biased estimates of the main genetic effect (i.e., the genetic effect in a population of mean age=0) genetic effects change with age. Correctly specified meta-regression can provide unbiased estimates of the main and age-varying genetic effects using summary level data, with a large number of studies covering a range of ages.

## Data Availability

The code used in this simulation study will become available at publication. All data produced in the present work are contained in the manuscript.

## DATA AVAILABILITY STATEMENT

The code used in this simulation study is available at (*link will become available at publication*). Summary data used in the applied example can be found in the original publication (10.1371/journal.pgen.1001307).

## AUTHOR CONTRIBUTIONS

PP, JPTH, TTM and KT contributed to the concept and design of the study. PP carried out all analyses and wrote the manuscript. All authors contributed to interpretation of the results and made critical revisions of the manuscript.

## ACKNOWLEDGEMENTS

We are grateful to Dr. Apostolos Gkatzionis for helpful comments on an earlier draft of the manuscript. No funders or people acknowledged influenced the study design or interpretation of results. The reviews expressed in this paper are those of the authors and not necessarily any funders or people acknowledged.

