## Supplementary Note for "Meta-regression of Genome-Wide Association Studies to estimate age-varying genetic effects"

**SUPPLEMNTARY NOTES**

**Supplementary Note S1.** Estimation of combined genetic effect in fixed-effect and random-effects meta-analysis.

Both fixed-effect and random-effects meta-analyses provide estimates of the combined genetic effect of a given SNP across all studies ($\hat{\beta}_{SNP}$) and the between study variability ($\hat{\tau}^{2}$). The combined genetic effect is a weighted mean of the genetic effects across all studies:

$$\hat{\beta}_{SNP}=\frac{\sum_{j=1}^{n} W_{j}\times\hat{\beta}_{1j}}{\sum_{j=1}^{n} W_{j}}$$

where $W_{j}$ is the weight assigned to each study and $\hat{\beta}_{1j}$ is the estimated effect for given SNP in each study. The fixed-effect model has only one source of variance (within-study variance due to sampling error), while the random-effects model has two sources of variance (within- and between-study variance). Therefore, the study weights are $W_{j}=\frac{1}{{\hat{SE(\beta_{1j})}}^{2}}$ and $W_{j}=\frac{1}{{\hat{SE(\beta_{1j})}}^{2}+ \tau^{2}}$ for fixed-effect and random-effects models, respectively.
